# Detecting Monogenic Obesity: A Systematic Exome-Wide Workup of Over 500 Individuals

**DOI:** 10.1101/2025.02.19.25322216

**Authors:** Robert Künzel, Helene Faust, Linnaeus Bundalian, Matthias Blüher, Mariami Jasaszwili, Anna Kirstein, Albrecht Kobelt, Antje Körner, Denny Popp, Eric Wenzel, Rami Abou Jamra, Johannes R. Lemke, Torsten Schöneberg, Robert Stein, Antje Garten, Diana Le Duc

**Affiliations:** University of Leipzig Medical Center, Institute of Human Genetics, Leipzig, Germany; University of Leipzig Medical Center, Medical Department III—Endocrinology, Nephrology, Rheumatology, Leipzig, Germany; Helmholtz Institute for Metabolic, Obesity and Vascular Research (HI-MAG) of the Helmholtz Zentrum München at the University of Leipzig and University Hospital Leipzig, Leipzig, Germany; University Hospital for Children and Adolescents, Pediatric Research Center, Leipzig, Germany; Chemnitz Clinic, Diagnostic Center GmbH MVZ, Practice for Human Genetics, Chemnitz, Germany; LIFE Child, LIFE Leipzig Research Center for Civilization Diseases, University of Leipzig, Germany; German Center for Child and Adolescent Health (DZKJ), Leipzig/Dresden partner site, Leipzig, Germany; Rudolf Schönheimer Institute of Biochemistry, Medical Faculty, University of Leipzig, Leipzig, Germany

## Abstract

**Background/Objectives:** Obesity poses a major public health concern. Although BMI heritability is estimated at 40–80%, genetic diagnostics remain challenging. This study aims to (*i*) assess the diagnostic yield of monogenic obesity in a large patient sample using exome-wide data, (*ii*) identify predictors to improve genetic testing criteria, and (*iii*) evaluate whether the identified genes are included in public obesity gene panels.

**Subjects/Methods:** We reviewed the genetic test results of 521 patients with obesity. 84.7% underwent whole-exome analysis, 15.3% were analyzed using a multi-thousand gene panel.

**Results:** Monogenic obesity was diagnosed in 5.8% of patients, while 7.1% carried a potentially obesogenic variant. Diagnostic yield was higher in children (6.3%) and patients with syndromic obesity (7.0%). Surprisingly, diagnostic yield was lower in severe obesity cases. 40% of patients with monogenic obesity carried variants in genes not included in current obesity panels.

**Conclusion:** Overall, 12.9% of patients had monogenic obesity or a potentially obesogenic variant. These findings suggest that genetic testing should not be limited to patients with extreme obesity. Current obesity panels miss crucial syndromic genes, demonstrating a need for more comprehensive panels and the superiority of whole exome sequencing in obesity.

## INTRODUCTION

Obesity, defined as a body mass index (BMI) over 30 kg/m² in adults (1) and above the 97LL percentile in children (2, 3), constitutes a profound and escalating global health challenge. Since 1990, obesity rates have more than doubled in adults and quadrupled in children, resulting in over one billion people worldwide living with obesity (4). The etiology of obesity is complex, arising from a confluence of environmental, behavioral, and genetic determinants (5). Among these, genetics is increasingly recognized for its critical role in influencing an individual’s susceptibility to obesity (6, 7). Notably, a systematic review of twin, family, and adoption studies concluded that BMI heritability ranges from 40−50% in the general population and rises to 80% in subpopulations with obesity, underscoring a substantial genetic component in obesity risk (8).

Obesity of genetic origin is classified into a polygenic and a monogenic form, although recent findings indicate a significant overlap between these two groups (9). Polygenic obesity arises from the cumulative impact of numerous genetic variants dispersed across various loci, with each variant showing only a minor association with the aggregate risk of obesity development (10). Conversely, monogenic obesity is characterized by a condition in which a solitary genetic variant significantly elevates an individual’s susceptibility to obesity, frequently resulting in severe obesity beginning at an early age (8). Estimates on the prevalence of monogenic obesity vary, depending on each study’s inclusion criteria and population. Considering only variants in known obesity genes classified as “pathogenic” and “likely pathogenic” according to the American College of Medical Genetics (ACMG) standards, diagnostic yields of 2.7% (11), 3.9% (12), to as high as 13% (13) have been reported in patients with obesity. This uncertainty emphasizes the need for additional research to provide a realistic insight into the genetic diagnostic yield of monogenic obesity.

Furthermore, monogenic obesity can be classified based on the presence or absence of developmental delay, intellectual disability, and/or dysmorphisms (DD/ID/D) (14). In recent years, pharmacological options for treating both these forms of monogenic obesity have begun to emerge (15), with setmelanotide being a notable example (16). However, as genetic diagnostics is expensive and not ubiquitously available, it is not part of routine screening for patients with obesity. Consequently, given that patients may benefit from a genetic diagnosis by receiving targeted treatment, there is a critical need for reliable predictors of genetic obesity to enhance diagnostic efficiency.

Most clinical research on monogenic obesity so far has focused solely on panels of selected obesity genes (11–13, 17–20). By examining only a predefined set of genes in the analysis, variants can be missed, especially in genes where an obesity association has only recently been described. Although sequencing costs decline, making whole-exome sequencing (WES) increasingly accessible, reports of patients with obesity who largely received WES in a diagnostic setting, including evaluation of copy number variants (CNVs), are currently missing. This gap poses a significant limitation in current studies related to monogenic obesity, highlighting the necessity for a broader investigative approach in this complex condition.

In this context, this exploratory study aims to (*i*) identify the diagnostic yield of monogenic obesity using exome-wide data in patients with syndromic and non-syndromic obesity. Additionally, it aims to (*ii*) determine traits predictive of a genetic diagnosis by phenotypically characterizing the cohort. Furthermore, this study aims to (*iii*) evaluate whether genes identified as associated with obesity in this cohort are also included in standard obesity gene panels.

## METHODS

### Patients

In order to identify patients with obesity who received WES or genetic diagnostics based on large clinical panels, we initially reviewed the results of individuals referred to the Institute of Human Genetics, University of Leipzig Medical Center, Germany, between 2016 and 2023 (*N* = 16 840). Patients were filtered by Human Phenotype Ontology terms assigned during presentation at our institute. In this study, we included 521 patients for whom obesity had been recorded, unless their reported BMI at the time of testing did not align with established obesity definitions. For the fourteen patients who underwent bariatric surgery, pre-surgery BMI was considered. A flowchart describing the cohort selection process is presented in Supplementary Figure 1.

### Patient Subgroups

Patients were divided into subgroups based on sex (male and female), age (adults and children, with children defined as those younger than 18 years at the time of testing), and the type of obesity (additional DD/ID/D or not). Furthermore, the severity of obesity was categorized: for adults, milder obesity was defined as a BMI of at least 30 but less than 40 kg/m², and severe obesity as a BMI of 40 kg/m² or higher. For children, milder obesity was classified as a BMI at or above the 97^th^ and below the 99.5^th^ percentile and severe obesity as a BMI at or above the 99.5^th^ percentile. Age- and sex-specific BMI percentiles for children were calculated using the German reference standards provided by Kromeyer-Hauschild *et al.* (2, 3).

### Sequencing Techniques

All individuals included in this cohort received next-generation sequencing (NGS)-based genetic diagnostics. The majority of probands, 84.5% (*n* = 440), received WES, using either a TWIST Human Core Exome kit (*n* = 257), TWIST Exome 2.0 (*n* = 147; TWIST Bioscience, San Francisco, USA), BGI Exome capture 59M (*n* = 26; BGI, Shenzhen, China), Nextera Exome Rapid Capture v1.2 (*n* = 7; Illumina, San Diego, USA) or Agilent Exome SureSelect v6 (*n* = 3; Agilent, Santa Clara, USA). Sequencing was performed on Illumina NovaSeq 6000 (TWIST, TWIST 2.0), BGISEQ-500 (BGI), or an Illumina NextSeq- or HighSeq platform (Nextera, Agilent). The remaining 15.3% of patients (*n* = 81) received genetic diagnostics based on the TruSight One Sequencing Panel (4 813 genes, Supplementary Table 1), using the TruSight Rapid Capture Kit and Illumina NextSeq550. CNV analysis was carried out based on microarray or NGS data.

### Variant Interpretation

Evaluation of variants was performed using the software tools Varvis and Varfeed (Limbus, Rostock, Germany). All variants were classified in accordance with ACMG criteria (21), as well as the Association for Clinical Genetic Science (ACGS) Best Practice Guidelines (22). Variants of unknown significance (VUS) were re-evaluated in selected patients strongly suspected of having genetic obesity. Based on the variants reported to referring physicians and to the patients, subjects were categorized into three groups regarding a monogenic obesity diagnosis: “solved”, “possibly solved” and “unsolved”. The criteria for categorizing these groups were as follows: individuals with variants classified as “pathogenic” or “likely pathogenic” in genes with an established association with obesity, as documented by the Online Mendelian Inheritance in Man (OMIM) (23) and/or GeneReviews (24) databases, were considered “solved”; patients who had VUS reported in recognized obesity genes, and those with a (likely) pathogenic variant in a gene with limited, but some documented evidence for a monogenic obesity association, were classified as “possibly solved”; all other individuals were designated “unsolved”.

### Panel Comparison

We investigated whether the obesity genes identified in this cohort were also included in publicly available obesity panels. To this end, we created one large panel consisting of all genes and loci listed in the obesity panels of two major sources for genetic panels, namely PanelApp Australia (25) and Genomics England PanelApp (26). These genes were then compared with the genes affected in patients with monogenic obesity in our cohort. The combined public panel consisted of the following 57 loci: *ACBD6, ADCY3, AKR1C2, ALMS1, ARL6, BBIP1, BBS1, BBS10, BBS12, BBS2, BBS4, BBS5, BBS7, BBS9, C8orf37, CEP164, CEP19, CEP290, CPE, DYRK1B, GNAS, HTR2C, IFT172, IFT27, IFT74, INPP5E, KIDINS220, KSR2, LEP, LEPR, LZTFL1, MAGEL2, MC4R, MKKS, MKS1, MRAP2, MYT1L, NR0B2, NTRK2, PCSK1, PGM2L1, PHF6, PHIP, POMC, PPARG, SCAPER, SDCCAG8, SH2B1, SIM1, TRIM32, TTC8, TUB, VPS13B, WDPCP*, 15q11q13 recurrent region (PWS/AS, BP1-BP3, Class 1) Loss, 15q11q13 recurrent region (PWS/AS, BP2-BP3, Class 2) Loss, 16p11.2 recurrent region (includes *SH2B1*, distal region, BP2-BP3) Loss.

### Statistical Analysis

Permutation tests were performed using R Studio (version 2024.04.0, Build 735) with the R programming language (version 4.4.0). A significance level of 0.05 was used to determine statistical significance.

### Use of Large Language Models

ChatGPT (Open AI, San Francisco, California, USA) was utilized to refine the clarity of writing in this manuscript. The model was employed selectively to enhance readability and coherence, without altering the substantive content or scientific rigor of the work.

## RESULTS

### Patient Characteristics

This study included 521 individuals with obesity, comprising 76% children (*n* = 396) and 24% adults (*n* = 125). Slightly more than half of the patients were male (54.9%). 57.4% of the patients (*n* = 299) showed obesity with additional developmental delay, intellectual disability, and/or dysmorphisms (DD/ID/D), while the remaining 42.6% (*n* = 222) had obesity without these additional features. Regarding the severity of obesity, 25.5% of the individuals (*n* = 133) showed a milder form of obesity and 51.1% of the patients (*n* = 266) had severe obesity. For the remaining 23.4% (*n* = 122), no BMI data was recorded beyond the diagnosis of obesity. For detailed patient characteristics, see Supplementary Table 2.

### Diagnostic Yield

In this study, 5.8% of the patient cases (*n* = 30) were classified as solved and therefore received a monogenic obesity diagnosis. Among these individuals, eight had a (likely) pathogenic variant in *MC4R*; five had a 16p11.2 microdeletion. Three patients had loss-of-function variants in *SRRM2*, and another three carried variants in *PHIP*. A complete list of affected genes is provided in Figure 1A. Table 1A and 1B present patient information and exact variant localization.

**Fig. 1.**
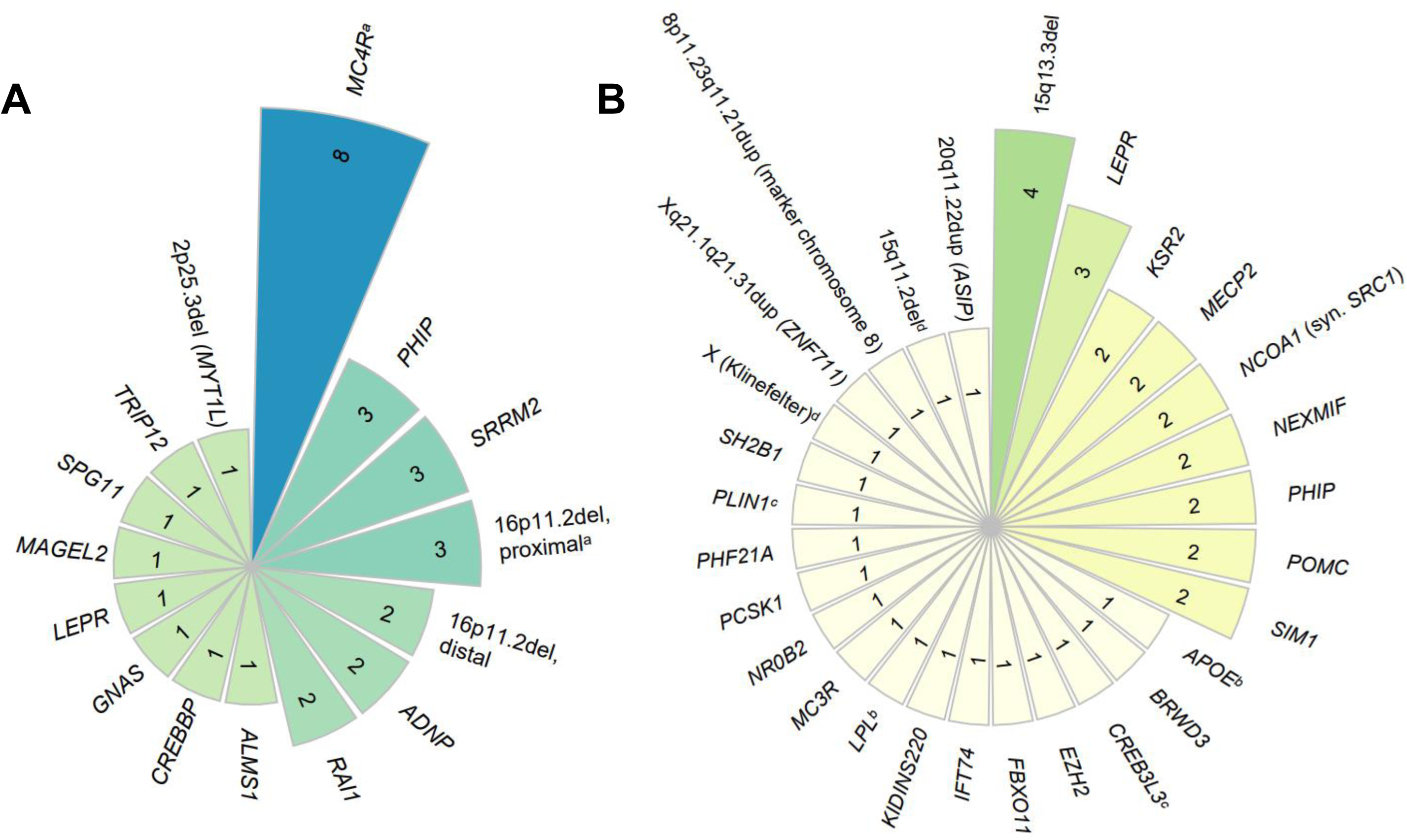
Number of cases per locus. This radial plot shows how many individuals carry a (possibly) obesogenic variant for each locus. **A.** Solved cases (L: patient with two affected loci). **B.** Possibly solved cases (L, L, L: patients with two affected loci).

An additional 7.1% of patients (*n* = 37) were considered possibly solved regarding monogenic obesity, with a potentially obesogenic variant identified in these individuals. A list of impacted genes can be seen in Figure 1B, and a list of patients and variants is available in Table 2A and 2B.

In total, 12.9% (*n* = 67) of the patients in this study carried a variant with a definite or suspected monogenic obesity association.

### Predictive Genetic Obesity Traits

Diagnostic yield was higher in children compared to adults, at 6.3% and 4.0%, respectively. Male and female patients exhibited similar rates of monogenic obesity (5.9% *vs.* 5.5%). However, patients with obesity and additional DD/ID/D were more likely to have a monogenic obesity cause compared to those without these additional features (7.0% *vs.* 4.1%). Notably, a higher BMI did not appear to increase the likelihood of a genetic obesity diagnosis. Among patients with severe obesity, 5.6% received a genetic obesity diagnosis, compared to 9% of individuals with milder obesity. A permutation analysis showed no significant statistical differences between groups for any of these results. An overview of diagnostic yields in selected subgroups can be found in Figure 2, an extensive list of subgroup yields including statistical results is presented in Supplementary Table 3. Supplementary Table 4 lists genes affected in patients with and without additional syndromic features.

**Fig. 2:**
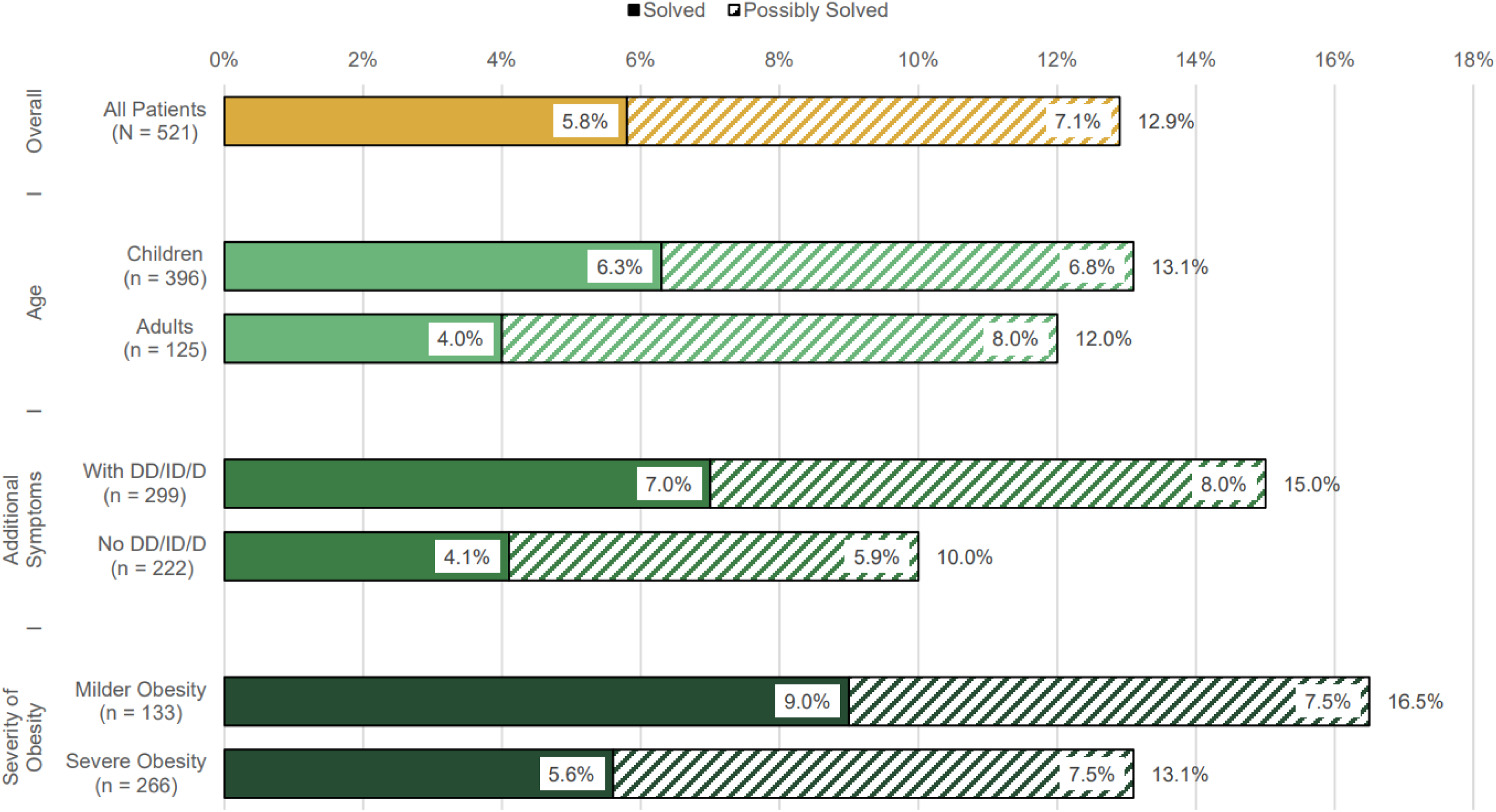
Diagnostic yield in selected subgroups. This bar chart demonstrates the percentage of solved and possibly solved cases inside several subgroups. Children, individuals with syndromic obesity and patients with milder obesity showed higher genetic diagnostic yields. (DD/ID/D: developmental delay, intellectual disability and/or dysmorphisms)

### Comparison with Obesity Panels

Among all solved monogenic obesity cases from this study, only 60% (18/30) of the variants were in genes also listed in the combined obesity panel described above. This discrepancy primarily stems from missing genes associated with syndromic obesity. In patients without additional syndromic features, around 90% (8/9) of affected loci were included in the obesity panel. Conversely, in patients with additional DD/ID/D, only approximately 50% (10/21) of solved obesity cases could be explained solely by genes included in the obesity panel. Results are presented in Figures 3A and 3B, a detailed overview can be found in Supplementary Table 4.

**Fig. 3:**
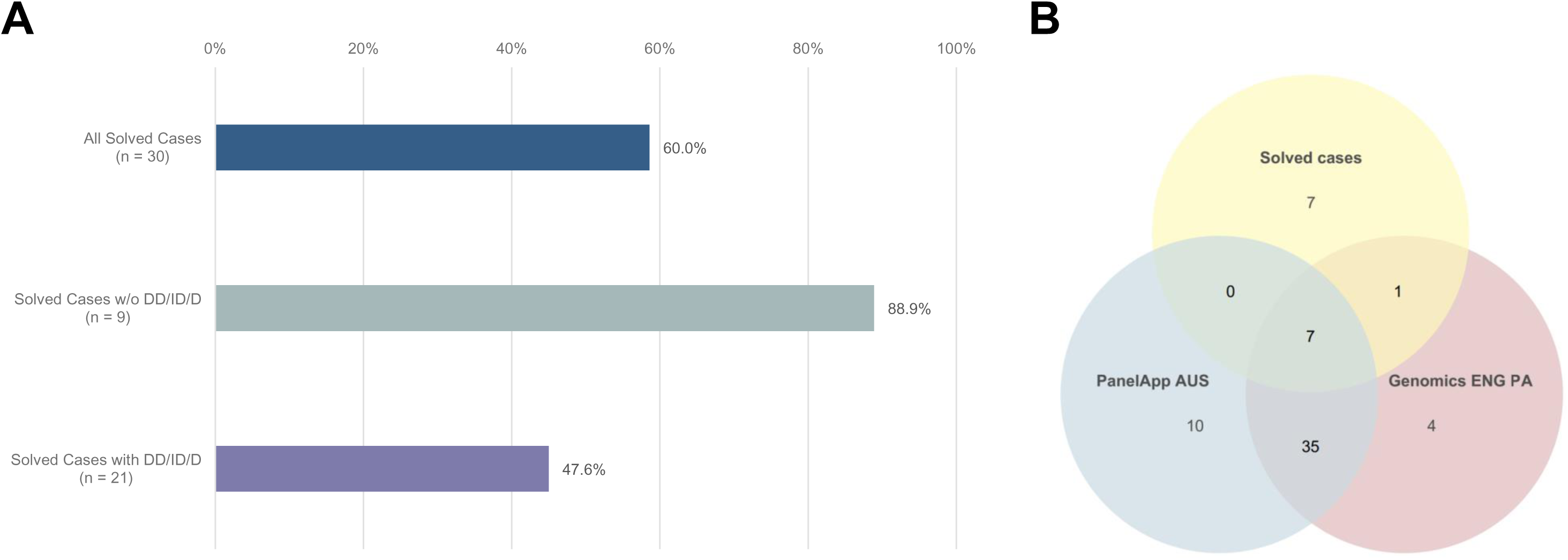
Comparison of public obesity panel genes and genes affected in patients with monogenic obesity in this cohort. **A** shows the percentage of solved cases that could be identified using only obesity panels. It distinguishes between patients with non-syndromic and syndromic monogenic obesity. **B** shows a Venn diagram demonstrating the overlap between genes included in public obesity panels and genes affected in solved obesity cases in our cohort. (DD/ID/D: developmental delay, intellectual disability and/or dysmorphisms; PA: PanelApp; w/o: without)

## DISCUSSION

This large retrospective study analyzed the genetic test results of 521 patients with obesity who were referred to our institute for obesity or other diagnoses, most of whom underwent WES. Overall, 5.8% of patients (*n* = 30) received a monogenic obesity diagnosis, while a further 7.1% (*n* = 37) carried a possibly obesogenic variant. A higher diagnostic yield was found in children and individuals with additional DD/ID/D. Counterintuitively, people with severe obesity exhibited a lower diagnostic yield compared to those with milder obesity. Lastly, WES identified several genetic obesity causes that would have been missed by panel diagnostics.

The first aim of this study was to report the genetic diagnostic yield of obesity. With 5.8%, the diagnostic yield in this patient sample was comparable to that of other studies, with some reporting lower (12, 17) and others reporting higher results (13, 18). Several systematic reviews support this outcome, estimating the prevalence of monogenic obesity at 5−10% in cohorts of patients with obesity (8, 27). As the patients in this cohort primarily received broad WES instead of selected panel diagnostics, we had assumed that a higher number of genetic causes would be identified, which was, however, not the case. This lack of increase despite more extensive testing could potentially be explained by (*i*) the cohort selection process, (*ii*) the interpretation of variants in this study, and (*iii*) the underreporting of obesity as a comorbidity. Firstly, all patients with obesity who received genetic testing at the Institute of Human Genetics, Leipzig, were included in this analysis, regardless of primary indication. Therefore, in some cases, the initial diagnostic focus could have been elsewhere (e.g., epilepsy, DD), and clinical features prompting the clinician to order genetic testing did not necessarily need to suggest a genetic cause for the obesity as well (e.g., lack of early onset). Secondly, a distinction was made between patients with definitive and possible genetic obesity. Due to the strict criteria applied for genetic variant interpretation, it is possible that some variants classified as VUS may indeed have a profound obesogenic effect. As genetic variant evaluation improves with more available data, the classification of some variants may change in the future; including the possibly solved cases, the diagnostic yield in this cohort could potentially increase to 12.9% (*n* = 67). For variants with unclear implications, proof of loss of function should be sought by functional studies to guide treatment decisions, as exemplified by patients with variants in the leptin receptor *(LEPR*) (28). A third factor that may alter the diagnostic yield in this study is incomplete documentation of obesity in patients in medical records. Obesity was reported in less than 5% of all individuals in the in-house database we analyzed (*N* = 16 840, of which 12 291 were index patients comprehensively phenotyped). This percentage significantly differs from the obesity prevalence of approximately 19% in the German population (29). While this discrepancy partly reflects the younger age distribution of our cohort, it also indicates underreporting of obesity when ordering genetic testing, especially if other, more severe symptoms are present. A recent study on the prevalence of comorbidities in individuals with neurodevelopmental delay found that clinical synopses on OMIM are missing about one-third of significantly enriched clinical features (30). Although the authors did not explicitly mention obesity as one of those underreported comorbidities, the issue of obesity not having been documented when genetic testing was ordered was repeatedly encountered during this study. As the evaluation of genetic variants heavily relies on correct and comprehensive phenotyping (31, 32), this underscores the necessity to thoroughly report on obesity.

In our study of patients with monogenic obesity, *MC4R* emerged as the most commonly affected gene. Eight patients carried a relevant variant in this gene, representing 1.5% of the total cohort and 26.7% of those with monogenic obesity. This result is supported by the current literature, which has repeatedly shown *MC4R* to be the most frequent monogenic obesity cause (12, 13, 27, 33). Additionally, five patients exhibited a 16p11.2 microdeletion, with three proximal and two distal deletions. Both forms are associated with obesity (34, 35), while recent research suggests higher obesity rates in patients with the 220 kbp distal deletion (36). This is in part due to this region encompassing the gene *SH2B1*, which affects central leptin-melanocortin and insulin signaling pathways (37). The third most frequently affected loci in patients with monogenic obesity in this cohort were *PHIP* and *SRRM2*, each impacted in three individuals. Variants in *PHIP* cause Chung-Jansen syndrome, linked to DD/ID/D and obesity (38–40). This established gene-phenotype association enabled the reclassification of the variant of patient #16 (Table 1A) from VUS to likely pathogenic, confirming the diagnosis of Chung-Jansen syndrome. The initial assessment took place in 2016, when data on the effect of *PHIP* variants were still scarce. This demonstrates the ongoing process of genetic variant evaluation improving with new research, again highlighting that future interpretations of variants in this study may change. Lastly, loss-of-function variants in *SRRM2* have been described by Cuinat *et al.* in 2022 to cause a neurodevelopmental disorder with facial dysmorphisms that is associated with overweight/obesity (41). Interestingly, patient #24 (Table 1A) showed severe obesity with a BMI above the 99.9^th^ percentile, but without DD/ID/D. This unusual phenotypic expression of *SRRM2* loss-of-function emphasizes the heterogeneity that can complicate the correct diagnosis of monogenic obesity. Adding to this, we excluded a girl with a pathogenic *SRRM2* variant because her BMI was just below the obesity threshold (96^th^ percentile). This illustrates that while the clear distinction between overweight and obesity may benefit research standardization, it can also oversimplify what is essentially a continuum rather than a binary categorization.

Furthermore, in an additional 7.1% of patients (*n* = 37), possibly obesogenic variants were identified, warranting further research to investigate their relevance. Patient #51 (Table 2A) had a heterozygous, likely pathogenic variant in *PCSK1*, which encodes an enzyme critical to the central leptin-melanocortin pathway. While mono-allelic variants in *PCSK1* have been considered obesity-causing before (12, 13, 17, 42, 43), recent research questions the monogenic effect of heterozygous *PCSK1* variants on obesity (44), prompting us to assess these cases conservatively. Despite this, the truncating nature of the identified *PCSK1* variant suggests a potential loss of enzyme function, increasing the likelihood that it could cause obesity even in a heterozygous state (44, 45). Similarly, patients #55 and #56 (Table 2A) carried heterozygous variants in *POMC*. Recent studies suggest that these variants only slightly increase BMI, questioning their relevance in monogenic obesity (46). Concurrently, an ongoing trial (EMANATE, RM-493-035) is investigating the effect of setmelanotide on patients with suspected genetic obesity who carry variants in genes involved in the *MC4R* pathway. For example, patients with heterozygous, likely pathogenic variants in *PCSK1* or *POMC* are eligible for inclusion. Additionally, we identified two patients (patients #46 and #47, Table 2A) with a variant in *NCOA1* (47). Although this variant is classified as VUS, the patients fulfil the trial’s inclusion criteria. This highlights a gap between genetic diagnostics and potential therapeutic consequences, demonstrating that patients could receive targeted treatment even without a definitive genetic diagnosis.

The second aim of this study was to identify predictors for a genetic obesity diagnosis. Several characteristics indicated a higher likelihood of a monogenic obesity diagnosis, though none of these changes were statistically significant. This aligns with the findings of Tamaroff *et al*., who also did not determine any significant predictive parameters (19). The diagnostic yield was higher in children compared to adults, consistent with the report by Kleinendorst *et al*. (12). Providing all children with obesity access to genetic testing could be beneficial, given the potential for personalized treatment (15, 16, 48) and psychological benefits, such as relief from self-blame (49). However, pathogenic variants have also been identified in adults with obesity, which argues against excluding them from genetic testing, as highlighted by Tamaroff *et al*. (19). In our study, 7.0% of patients with additional DD/ID/D (*n* = 21) received a genetic obesity diagnosis, indicating that genetic obesity causes should be considered even when obesity is present only as an additional symptom. Interestingly, the diagnostic yield was not increased in patients with severe obesity. In fact, patients with milder obesity received a genetic diagnosis more frequently. Similarly, Kleinendorst *et al*. (12, 13) and Tamaroff *et al*. (19) did not identify any significant BMI differences in individuals with and without relevant variants. This suggests that genetic testing should not be limited to cases of extreme obesity.

The third insight of this study is that large public obesity panels lack several obesity genes affected in this cohort, particularly in patients with additional DD/ID/D. Although PanelApp Australia explicitly includes genes associated with syndromic obesity (25), the evaluated panels only focus on severe early-onset obesity, thereby excluding genes associated with milder or later-onset phenotypes. As discussed, patient #24 (Table 1A) in this study carried a variant in *SRRM2*, which typically causes an intellectual developmental disorder and obesity (41). However, the patient did not exhibit ID and would have been missed by obesity panel analysis alone. Likewise, Kleinendorst *et al*. identified six patients with a genetic obesity disorder typically associated with ID who did not have ID (13). Variants in patients without fully penetrant phenotypes are at risk of escaping evaluation if panels do not consider incomplete phenotypic expressivity of syndromic obesity.

In contrast, patient #6 (*LEPR*) and patients #11 and #14 (*MC4R*) carried pathogenic variants in genes typically associated with isolated obesity (14), but exhibited additional syndromic features (Table 1A). It is suspected that they harbor further pathogenic variants in DD-related genes, though none were identified in the analysis. This suggests that while individuals with syndromic obesity-associated variants can present with isolated obesity, the reverse is also possible. Consequently, extensive testing is needed to accurately diagnose these complex cases.

Moreover, ongoing research continues to expand the list of genes that determine body weight. For example, the rhodopsin-like G protein-coupled receptors (GPCRs) NPY2R and NPFFR2 (50), as well as the adhesion GPCR latrophilin 1 (ADGRL1/LPHN1) (51), significantly contribute to body weight regulation. Their relevance in causing obesity has been demonstrated at least in mice. A diagnostic approach using current panels might have missed such genes and their potential association with a human phenotype. This highlights the need for more comprehensive obesity panels and demonstrates the superiority of WES in obesity diagnostics.

## STRENGTHS AND LIMITATIONS

This study benefits from its large sample size of over 500 participants. Given the relatively low prevalence of monogenic obesity, this size allows for a more comprehensive analysis of the data and enhances the reliability of the findings. Moreover, patients in this cohort were not selected for diagnostics based on suspicion of genetically determined obesity. Thus, compared to other studies, the present cohort is less biased towards testing patients who strictly meet clinical criteria for monogenic obesity (such as hyperphagia or extreme BMI) and contributes to a broad and comprehensive perspective on the genetic causes of obesity. Most patients were analyzed using WES, which offers more extensive diagnostic capabilities compared to panel analyses. Variants were classified in accordance with ACMG and ACGS criteria, strengthening the significance and reproducibility of the results.

On the other hand, this study is limited by its retrospective approach. As discussed, obesity was likely not reported consistently, leading to several patients with obesity being missed by this analysis. This study was conducted using a clinical sample at a single institution, which may limit the generalizability of the findings to other populations and settings.

## CONCLUSION

In this study of patients with obesity, 12.9% received either a definitive monogenic obesity diagnosis (5.8%) or carried a possibly obesogenic variant (7.1%). Although differences were not statistically significant, genetic diagnostic yield was higher in children and patients with additional DD/ID/D, suggesting a low threshold for genetic obesity testing in these groups. Diagnosis of genetic obesity did not correlate with higher BMI, suggesting that genetic testing should not be limited to cases of extreme obesity. Obesity panels would have missed 40% of patients with monogenic obesity in this cohort, primarily due to the incomplete inclusion of genes associated with syndromic obesity. Consequently, more comprehensive obesity panels are needed. To improve future diagnostic results, obesity should be reported consistently when ordering genetic testing. In conclusion, given that over one billion people globally are living with obesity, the possibility of a genetic origin should not be dismissed. Millions may have a monogenic obesity cause and could potentially benefit from targeted treatment.

## Supporting information

All Supplements (except for Table 1)

## ACKNOWLEDGMENTS

We would like to thank all patients included in this study, as well as the external clinicians and scientists involved in collaborating with the Institute of Human Genetics Leipzig, in particular Emanuele Coci (Magdeburg), Matthias Drechsler (Düsseldorf), Wiebke Fenske (Bochum), Monika Kautza-Lucht (UK Schleswig-Holstein), Nils Rahner (Bonn) and Ina Schanze (Magdeburg). RS acknowledges support through the Medical Faculty, University of Leipzig (“Nachwuchsförderprogramm”), and the German Diabetes Association (DDG, “Allgemeine Projektförderung”).

## AUTHOR CONTRIBUTIONS

R.K. conception and design of the study, data extraction, analysis and interpretation, drafting and revision of the manuscript. D.L.D. and A.G.: conception and design of the study, supervision of data collection and interpretation, drafting and revision of the manuscript, funding acquisition. H.F., L.B., M.J., A.Ki.: conception of the study, data interpretation, drafting and revision of the manuscript. M.B., T.S., R.S., A.Kö., A.Kob., E.W.: patient coordination, interpretation of results, manuscript revision. R.A.J., J.L., D.P.: genetic diagnosis coordination, data analysis and interpretation, manuscript revision.

## DATA AVAILABILITY

The datasets generated during and/or analysed during the current study are available from the corresponding author on reasonable request.

## ETHICAL APPROVAL

This study was approved and monitored by the Ethics Committee of the University of Leipzig, Germany (224/16-ek and 402/16-ek) and was conducted in concordance to the declaration of Helsinki. All families provided written informed consent for genetic testing. Institutional ethics approval was not required if testing was part of routine clinical care.

---

A. SNVs in solved cases
| Patient† | Age group‡ | Sex | BMI (kg/m2) | BMI percentile (children) | BMI SDS (children) |
| --- | --- | --- | --- | --- | --- |
| 1 | Child | M | 19.8 | 97 | 1.91 |
| 2 | Adult | M | 30.2 |  |  |
| 3 | Child | F | 24.4 | 100 | 3.78 |
| 4 | Child | M | 25.6 | 97.8 | 2.03 |
| 5 | Child | M | - | - | - |
| 6 | Child | M | 57.9 | 100 | 4.23 |
| 7 | Child | M | 32.3 | 99.4 | 2.51 |
| 8 | Child | M | 48.3 | 100 | 3.44 |
| 9<sup>a</sup> | Adult | F | 67.5 |  |  |
| 10 | Child | F | 25.8 | 99.9 | 3.16 |
| 11 | Child | M | 32.3 | 99.5 | 2.56 |
| 12 | Child | F | 47 | 100 | 4.23 |
| 13 | Child | M | 22.6 | 98 | 2.04 |
| 14 | Child | F | 29.5 | 99.7 | 2.76 |
| 15 | Child | F | 26.8 | 99.7 | 2.75 |
| 16 | Child | F | 22.3 | 99.8 | 2.87 |
| 17 | Child | F | 37.5 | 99.9 | 3.15 |
| 18 | Child | M | 21.3 | 97.9 | 2.04 |
| 19 | Child | M | - | - | - |
| 20 | Child | F | 31.7 | 99.4 | 2.56 |
| 21 | Adult | F | 32 |  |  |
| 22 | Adult | M | - |  |  |
| 23 | Child | M | 23.9 | 98.2 | 2.1 |
| 24 | Child | M | 22.6 | 99.9 | 3.26 |
| 25 | Child | M | 32 | 98.8 | 2.27 |

B. CNVs in solved cases
| Patient† | Age group‡ | Sex | BMI (kg/m2) | BMI percentile (children) | BMI SDS (children) |
| --- | --- | --- | --- | --- | --- |
| 26 | Child | F | 35.1 | 100 | 5.25 |
| 9<sup>a</sup> | Adult | F | 67.5 |  |  |
| 27 | Child | M | 31.9 | 98.8 | 2.3 |
| 28 | Child | M | 33.6 | 99.7 | 2.73 |
| 29 | Adult | F | 34.5 |  |  |
| 30 | Child | F | 31.7 | 99.5 | 2.57 |

AD autosomal dominant, AR autosomal recessive, CNVs copy number variants, DD/ID/D dele
†: patients were numbered after the evaluation was completed; this had no impact on scientifi
‡: *child* younger than 18 years at the time of testing, *adult* 18 years or older
<sup>a</sup>: patient with variants in two loci

| Gene | OMIM phenotype | Inheritance |
| --- | --- | --- |
| *ADNP* | Helsmoortel-van der Aa syndrome (#615873) | AD |
| *ADNP* | Helsmoortel-van der Aa syndrome (#615873) | AD |
| *ALMS1* | Alstrom syndrome (#203800) | AR |
| *CREBBP* | Rubinstein-Taybi syndrome 1 (#180849) | AD |
| *GNAS* | Pseudohypoparathyroidism Ia (#103580) | AD |
| *LEPR* | Obesity, morbid, due to leptin receptor deficiency (#614963) | AR |
| *MAGEL2* | Schaaf-Yang syndrome (#615547) | AD |
| *MC4R* | Obesity (BMIQ20) (#618406) | AD |
| *MC4R* | Obesity (BMIQ20) (#618406) | AD |
| *MC4R* | Obesity (BMIQ20) (#618406) | AD |
| *MC4R* | Obesity (BMIQ20) (#618406) | AD |
| *MC4R* | Obesity (BMIQ20) (#618406) | AD |
| *MC4R* | Obesity (BMIQ20) (#618406) | AD |
| *MC4R* | Obesity (BMIQ20) (#618406) | AD |
| *MC4R* | Obesity (BMIQ20) (#618406) | AD |
| *PHIP* | Chung-Jansen syndrome (#617991) | AD |
| *PHIP* | Chung-Jansen syndrome (#617991) | AD |
| *PHIP* | Chung-Jansen syndrome (#617991) | AD |
| *RAI1* | Smith-Magenis syndrome (#182290) | AD |
| *RAI1* | Smith-Magenis syndrome (#182290) | AD |
| *SPG11* | Spastic paraplegia 11 (#604360) | AR |
| *SRRM2* | Intellectual developmental disorder, AD 72 (#620439) | AD |
| *SRRM2* | Intellectual developmental disorder, AD 72 (#620439) | AD |
| *SRRM2* | Intellectual developmental disorder, AD 72 (#620439) | AD |
| *TRIP12* | Intellectual developmental disorder, AD 49 (#617752) | AD |
| Locus | OMIM phenotype | CNV type |
| 2p25.3 (*MYT1L*) | Intellectual developmental disorder, AD 39 (#616521) | deletion |
| 16p11.2, proximal | Chromosome 16p11.2 deletion syndrome, 593 kbp (#611913) | deletion |
| 16p11.2, proximal | Chromosome 16p11.2 deletion syndrome, 593 kbp (#611913) | deletion |
| 16p11.2, proximal | Chromosome 16p11.2 deletion syndrome, 593 kbp (#611913) | deletion |
| 16p11.2, distal | Chromosome 16p11.2 deletion syndrome, 220 kbp (#613444) | deletion |
| 16p11.2, distal | Chromosome 16p11.2 deletion syndrome, 220 kbp (#613444) | deletion |

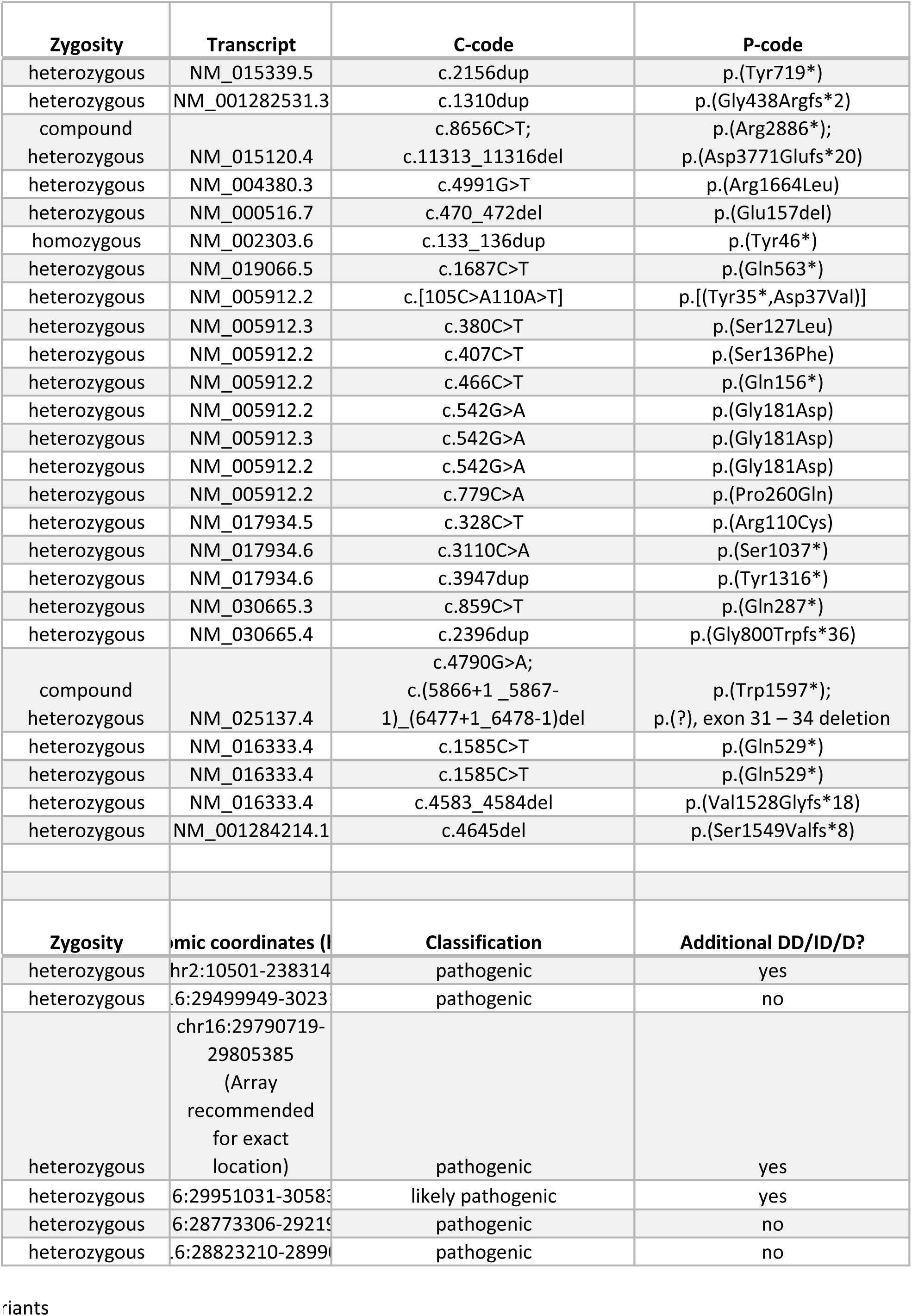

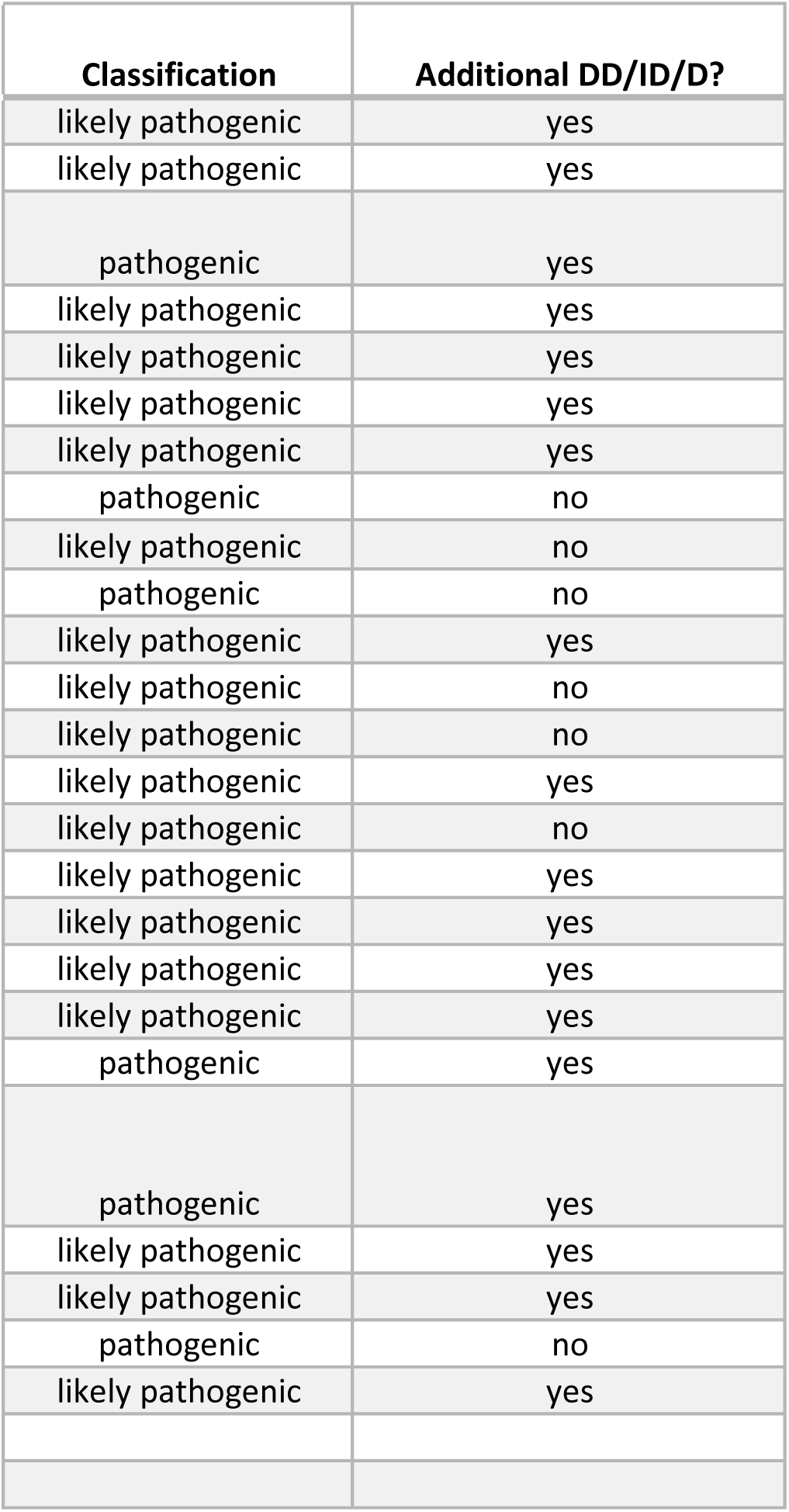

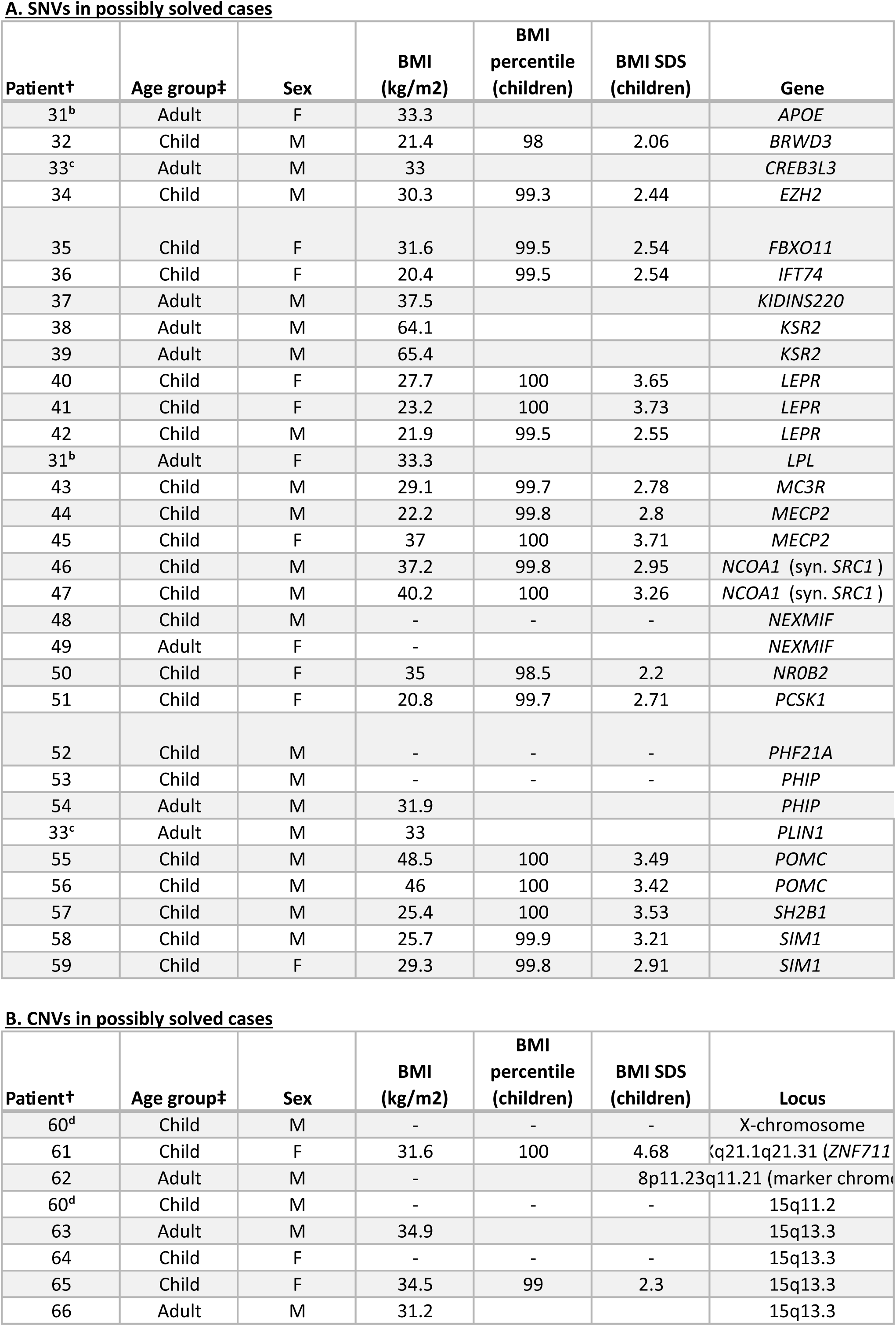
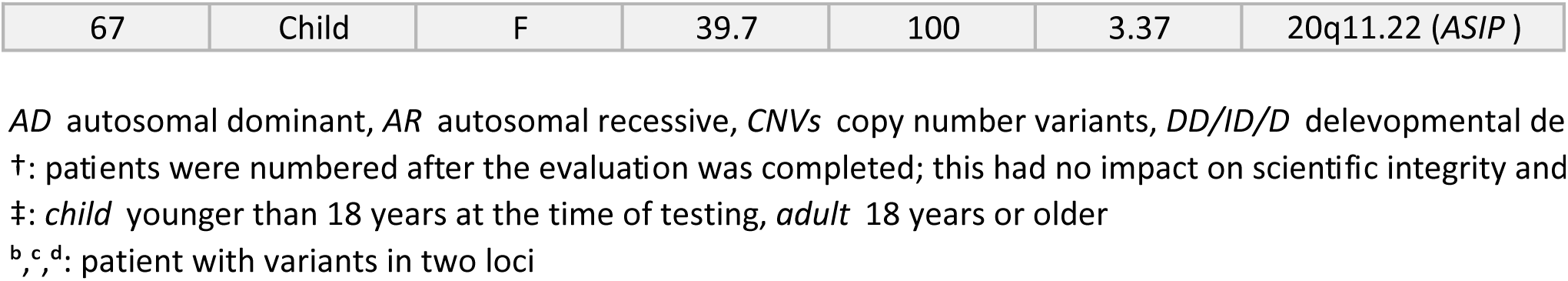

| OMIM phenotype | Inheritance |
| --- | --- |
| Hyperlipoproteinemia, type III (#617347) | - |
| Intellectual developmental disorder, X-L 93 (#300659) | XLR |
| Hypertriglyceridemia 2 (#619324) | AD |
| Weaver syndrome (#277590) | AD |
| Intellectual developmental disorder with dysmorphic facies and behavioral abnormalities (#618089) | AD |
| Bardet-Biedl syndrome 22 (#617119) | AR |
| Spastic paraplegia, intellectual disability, nystagmus, and obesity (#617296) | AD |
| - | - |
| - | - |
| Obesity, morbid, due to leptin receptor deficiency (#614963) | AR |
| Obesity, morbid, due to leptin receptor deficiency (#614963) | AR |
| Obesity, morbid, due to leptin receptor deficiency (#614963) | AR |
| Combined hyperlipidemia, familial (#144250) | AD |
| Obesity, severe, susceptibility to, BMIQ9 (#602025) | - |
| Intellectual developmental disorder, X-linked syndromic 13 (#300055) | XLR |
| Rett syndrome (#312750) | XLD |
| - | - |
| - | - |
| Intellectual developmental disorder, X-linked 98 (#300912) | XLD |
| Intellectual developmental disorder, X-linked 98 (#300912) | XLD |
| Obesity, mild, early-onset (#601665) | AD, AR, Mu |
| Obesity, susceptibility to, BMIQ12 (#612362) | - |
| Intellectual developmental disorder with behavioral abnormalities and craniofacial dysmorphism with or without seizures (#618725) | AD |
| Chung-Jansen syndrome (#617991) | AD |
| Chung-Jansen syndrome (#617991) | AD |
| Lipodystrophy, familial partial, type 4 (#613877) | AD |
| Obesity, early-onset, susceptibility to (#601665) | AD, AR, Mu |
| Obesity, early-onset, susceptibility to (#601665) | AD, AR, Mu |
| - | - |
| - | - |
| - | - |

| OMIM phenotype | CNV type |
| --- | --- |
| Klinefelter syndrome |  |
| Intellectual developmental disorder, XL 97 (#300803) | duplication |
| - | duplication |
| Chromosome 15q11.2 deletion syndrome (#615656) | deletion |
| Chromosome 15q13.3 microdeletion syndrome (#612001) | deletion |
| Chromosome 15q13.3 microdeletion syndrome (#612001) | deletion |
| Chromosome 15q13.3 microdeletion syndrome (#612001) | deletion |
| Chromosome 15q13.3 microdeletion syndrome (#612001) | deletion |
| - | duplication |

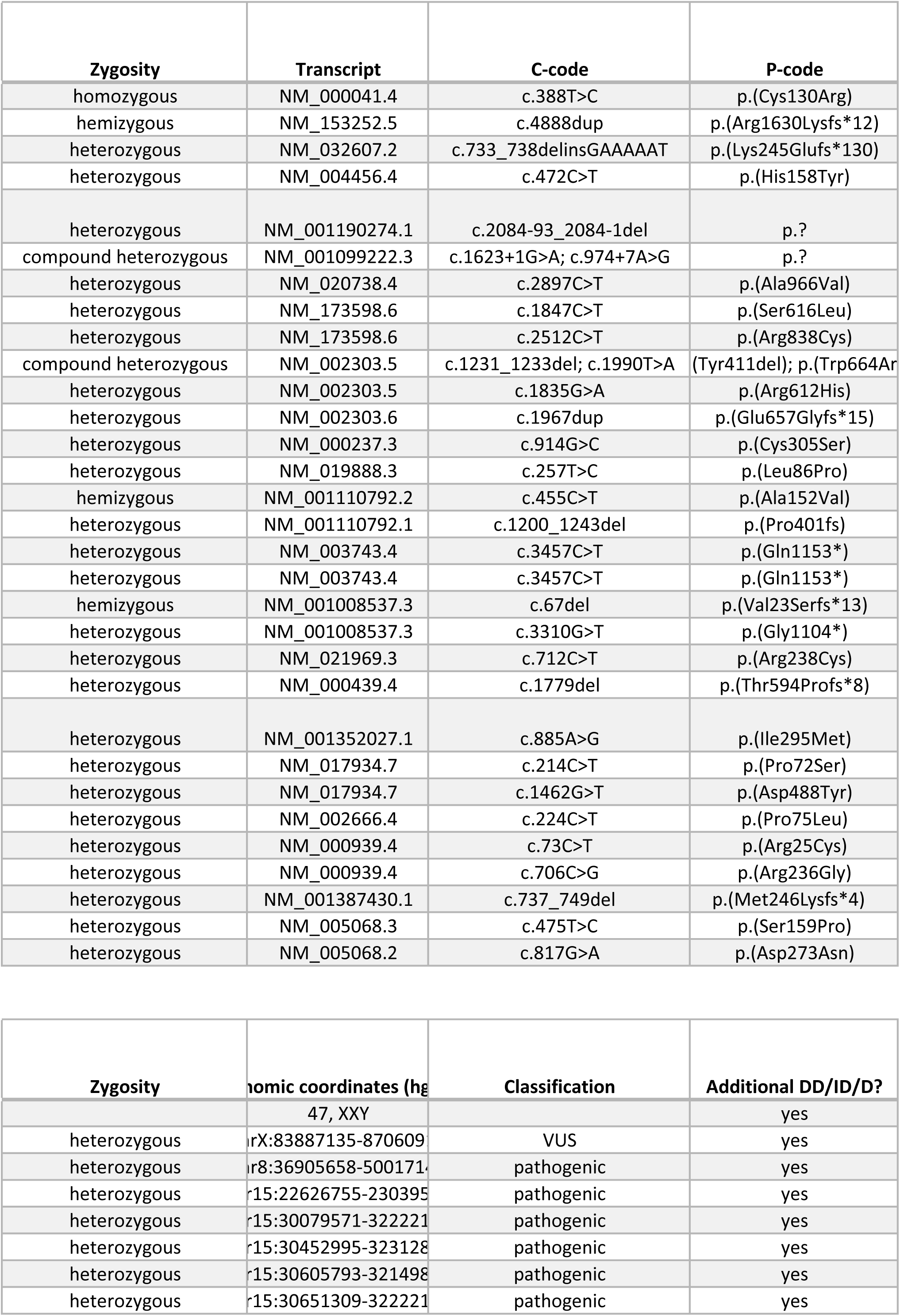
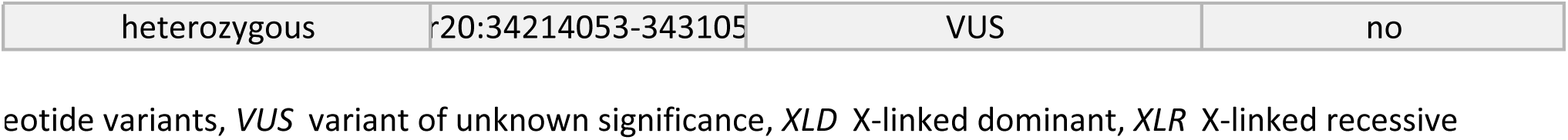

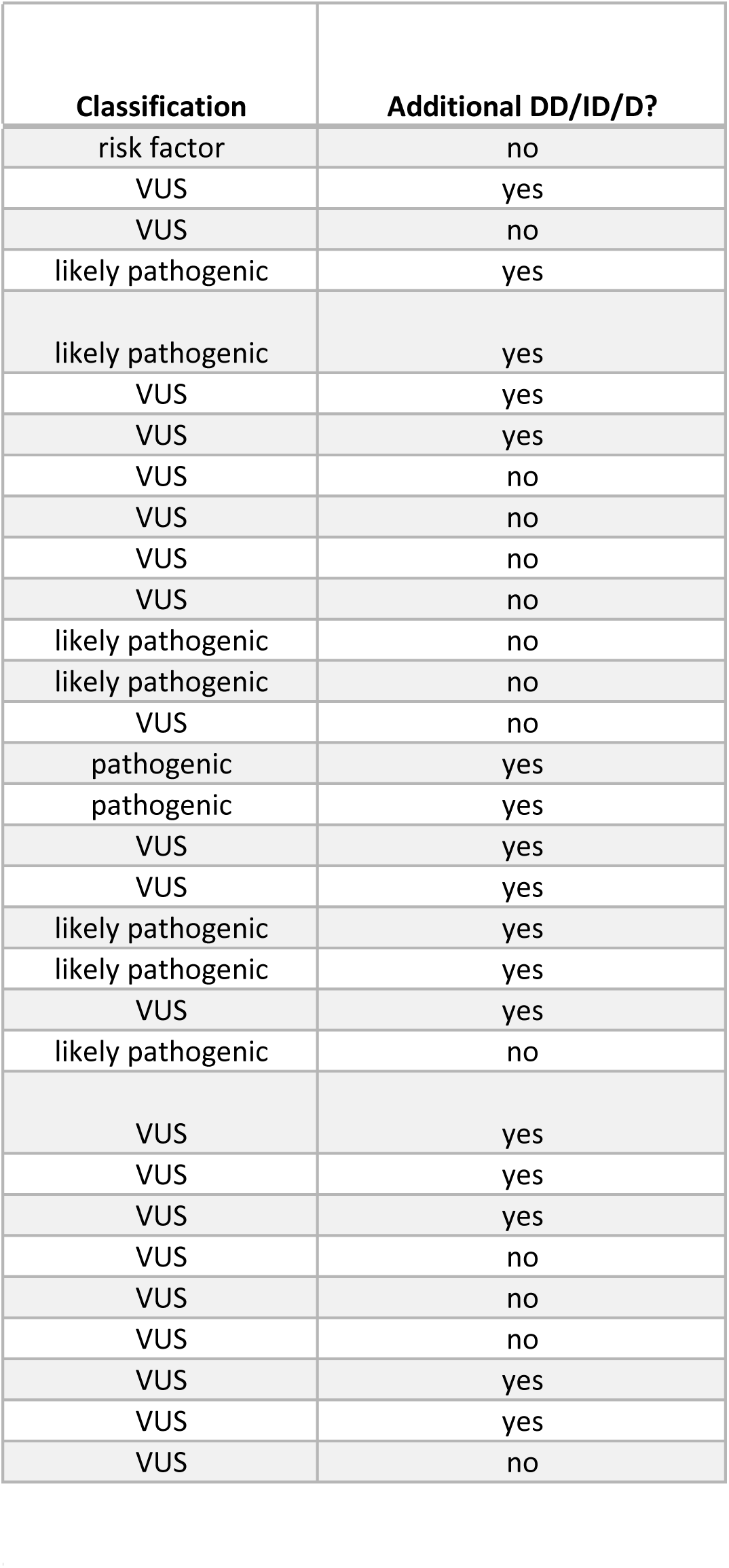

