## Supplementary material for "Detecting Monogenic Obesity: A Systematic Exome-Wide Workup of Over 500 Individuals": All Supplements (except for Table 1)

### <u>Supplementary Information</u>


- **Supp. Fig. 1:** Flowchart describing the study's cohort selection process: *p.2*
- **Supp. Table 1:** List of all genes included in the TruSight One Sequencing Panel (May 2014): *additional excel file*
- **Supp. Table 2:** Characteristics of patients in this study: *p.3*
- **Supp. Table 3:** Diagnostic yield in subgroups of patients: *p.4*
- **Supp. Table 4:** Comparison of affected genes in solved cases with obesity panel genes: *p.5*

Supplementary Figure 1 - Inclusion criteria

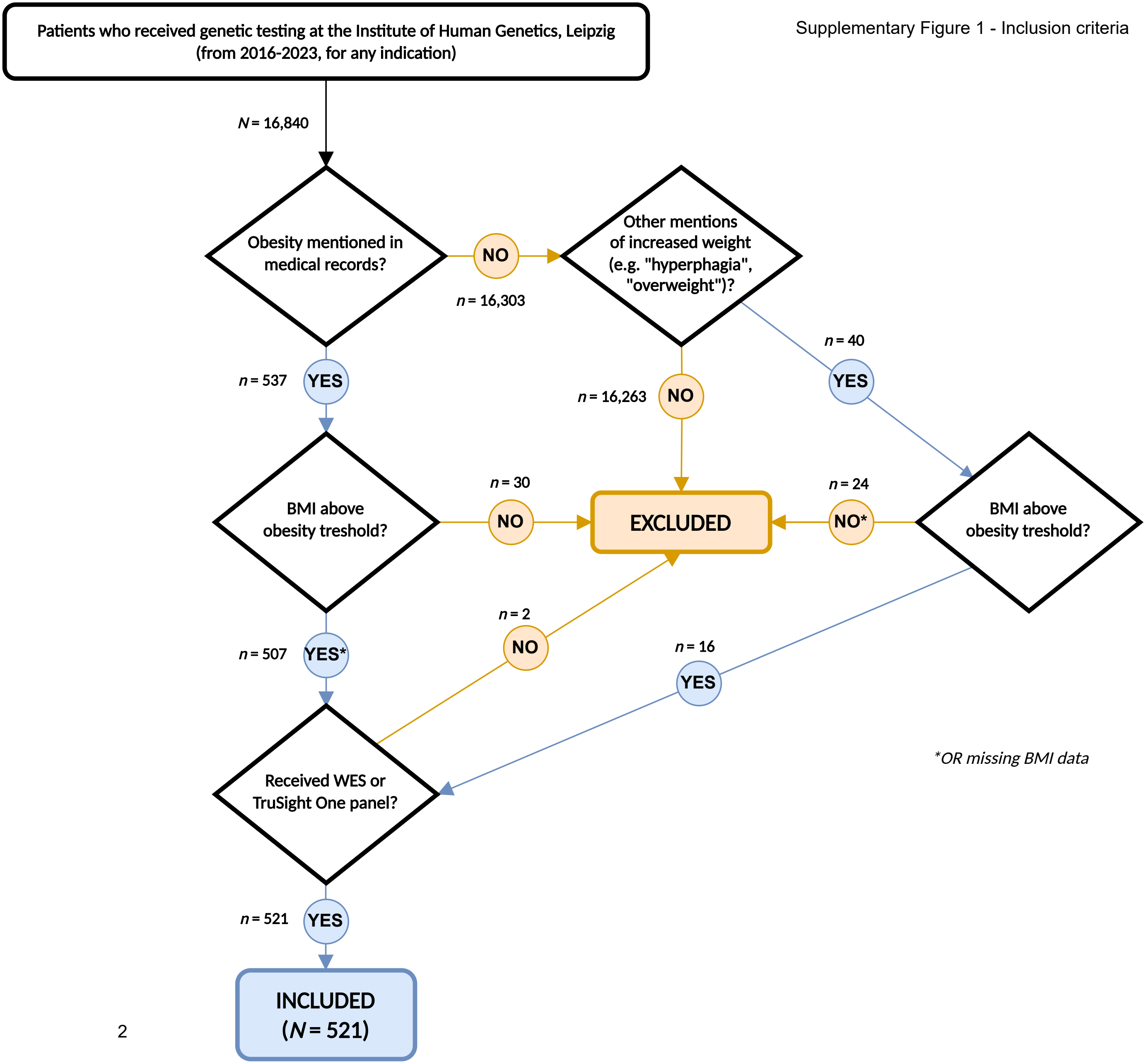

Supplementary Table 2 - Patient Characteristics

| | All Patients (N = 521) | | Solved (n = 30, 5.8%) | | Possibly solved (n = 37, 7.1%) | | Unsolved (n = 454, 87.1%) | |
|---|---|---|---|---|---|---|---|---|
| **Sex (n, %)** | | | | | | | | |
| Male | 286 | 54.9% | 17 | 56.7% | 23 | 62.2% | 246 | 54.2% |
| Female | 235 | 45.1% | 13 | 43.3% | 14 | 37.8% | 208 | 45.8% |
| **Age (n, %)** | | | | | | | | |
| Children | 396 | 76.0% | 25 | 83.3% | 27 | 73.0% | 344 | 75.8% |
| Adults | 125 | 24.0% | 5 | 16.7% | 10 | 27.0% | 110 | 24.2% |
| Mean, Median (years) | 16.3, 12.8 | | 13.8, 13.1 | | 17.3, 14.4 | | 16.4, 12.7 | |
| Range (years) | 0.3 – 79.8 | | 0.4 – 50.2 | | 1.6 – 57.6 | | 0.3 – 79.8 | |
| **Other Symptoms (n, %)** | | | | | | | | |
| Additional DD/ID/D | 299 | 57.4% | 21 | 70.0% | 24 | 64.9% | 254 | 55.9% |
| No DD/ID/D | 222 | 42.6% | 9 | 30.0% | 13 | 35.1% | 200 | 44.1% |
| **Severity of Obesity (n, %)** | | | | | | | | |
| Milder Obesity | 133 | 25.5% | 12 | 40.0% | 10 | 27.0% | 111 | 24.4% |
| Severe Obesity | 266 | 51.1% | 14 | 46.7% | 20 | 54.1% | 232 | 51.1% |
| BMI unknown | 122 | 23.4% | 3 | 10.0% | 7 | 18.9% | 112 | 24.7% |

| | Children (n = 396) | | Solved (n = 25) | | Possibly solved (n = 27) | | Unsolved (n = 344) | |
|---|---|---|---|---|---|---|---|---|
| **Sex (n, %)** | | | | | | | | |
| Male | 227 | 57.3% | 15 | 60.0% | 15 | 59.3% | 197 | 57.3% |
| Female | 169 | 42.7% | 10 | 40.0% | 12 | 44.4% | 147 | 42.7% |
| **Age (years)** | | | | | | | | |
| Mean, Median | 10.1, 10.2 | | 10.5, 11.4 | | 9.8, 8.8 | | 10.1, 10.3 | |
| Range | 0.3 – 17.9 | | 0.4 – 17.8 | | 1.6 – 17.3 | | 0.3 – 17.9 | |
| **Other Symptoms (n, %)** | | | | | | | | |
| Additional DD/ID/D | 221 | 55.8% | 18 | 72.0% | 18 | 66.7% | 185 | 53.8% |
| No DD/ID/D | 175 | 44.2% | 7 | 28.0% | 9 | 33.3% | 159 | 46.2% |
| **Severity of Obesity (n, %)** | | | | | | | | |
| Milder Obesity | 93 | 23.5% | 9 | 36.0% | 4 | 14.8% | 80 | 23.3% |
| Severe Obesity | 226 | 57.1% | 14 | 56.0% | 18 | 66.7% | 194 | 56.4% |
| BMI unknown | 77 | 19.4% | 2 | 8.0% | 5 | 18.5% | 70 | 20.3% |

| | Adults (n = 125) | | Solved (n = 5) | | Possibly solved (n = 10) | | Unsolved (n = 110) | |
|---|---|---|---|---|---|---|---|---|
| **Sex (n, %)** | | | | | | | | |
| Male | 59 | 47.2% | 2 | 40.0% | 8 | 80.0% | 49 | 44.5% |
| Female | 66 | 52.8% | 3 | 60.0% | 2 | 20.0% | 61 | 55.5% |
| **Age (years)** | | | | | | | | |
| Mean, Median | 36.2, 34.1 | | 31.0, 26.1 | | 37.5, 36.9 | | 36.3, 34.0 | |
| Range | 18 – 79.8 | | 18.1 – 50.2 | | 23.7 – 57.6 | | 18 – 79.8 | |
| **Other Symptoms (n, %)** | | | | | | | | |
| Additional DD/ID/D | 78 | 62.4% | 3 | 60.0% | 6 | 60.0% | 69 | 62.7% |
| No DD/ID/D | 47 | 37.6% | 2 | 40.0% | 4 | 40.0% | 41 | 37.3% |
| **Severity of Obesity (n, %)** | | | | | | | | |
| Milder Obesity | 40 | 32.0% | 3 | 60.0% | 6 | 60.0% | 31 | 28.2% |
| Severe Obesity | 40 | 32.0% | 1 | 20.0% | 2 | 20.0% | 37 | 33.6% |
| BMI unknown | 45 | 36.0% | 1 | 20.0% | 2 | 20.0% | 42 | 38.2% |

Supplementary Table 3 - Yield per Subgroup

| | All Patients (N = 521) | | Solved (n = 30) | | Permutation Test (p-value) | Possibly solved (n = 37) | | Unsolved (n = 454) | |
|---|---|---|---|---|---|---|---|---|---|
| **All Patients (N, %)** | 521 | 100% | 30 | 5.8% | | 37 | 7.1% | 454 | 87.1% |
| **Sex (n, %)** | | | | | | | | | |
| Male | 286 | 54.9% | 17 | 5.9% | 0.499 | 23 | 8.0% | 246 | 86.0% |
| Female | 235 | 45.1% | 13 | 5.5% | | 14 | 6.0% | 208 | 88.5% |
| **Age (n, %)** | | | | | | | | | |
| Children | 396 | 76.0% | 25 | 6.3% | 0.231 | 27 | 6.8% | 344 | 86.9% |
| Adults | 125 | 24.0% | 5 | 4.0% | | 10 | 8.0% | 110 | 88.0% |
| Mean, Median (years) | 16.3, 12.8 | | 13.8, 13.1 | | | 17.3, 14.4 | | 16.4, 12.7 | |
| Range (years) | 0.3 – 79.8 | | 0.4 – 50.2 | | | 1.6 – 57.6 | | 0.3 – 79.8 | |
| **Other Symptoms (n, %)** | | | | | | | | | |
| Additional DD/ID/D | 299 | 57.4% | 21 | 7.0% | 0.105 | 24 | 8.0% | 254 | 84.9% |
| No DD/ID/D | 222 | 42.6% | 9 | 4.1% | | 13 | 5.9% | 200 | 90.1% |
| **Severity of Obesity (n, %)** | | | | | | | | | |
| Milder Obesity | 133 | 25.5% | 12 | 9.0% | 0.147 | 10 | 7.5% | 111 | 83.5% |
| Severe Obesity | 266 | 51.1% | 15 | 5.6% | | 20 | 7.5% | 231 | 86.8% |
| BMI unknown | 122 | 23.4% | 3 | 2.5% | | 7 | 5.7% | 112 | 91.8% |

| | Children (n = 396) | | Solved (n = 25) | | Permutation Test (p-value) | Possibly solved (n = 27) | | Unsolved (n = 344) | |
|---|---|---|---|---|---|---|---|---|---|
| **All Children (n, %)** | 396 | 100.0% | 25 | 6.3% | | 27 | 6.8% | 344 | 86.9% |
| **Sex (n, %)** | | | | | | | | | |
| Male | 227 | 57.3% | 15 | 6.6% | 0.479 | 15 | 6.6% | 197 | 86.8% |
| Female | 169 | 42.7% | 10 | 5.9% | | 12 | 7.1% | 147 | 87.0% |
| **Age (years)** | | | | | | | | | |
| Mean, Median | 10.1, 10.2 | | 10.5, 11.4 | | | 9.8, 8.8 | | 10.1, 10.3 | |
| Range | 0.3 – 17.9 | | 0.4 – 17.8 | | | 1.6 – 17.3 | | 0.3 – 17.9 | |
| **Other Symptoms (n, %)** | | | | | | | | | |
| Additional DD/ID/D | 221 | 55.8% | 18 | 8.1% | 0.068 | 18 | 8.1% | 185 | 83.7% |
| No DD/ID/D | 175 | 44.2% | 7 | 4.0% | | 9 | 5.1% | 159 | 90.9% |
| **Severity of Obesity (n, %)** | | | | | | | | | |
| Milder Obesity | 93 | 23.5% | 9 | 9.7% | 0.195 | 4 | 4.3% | 80 | 86.0% |
| Severe Obesity | 226 | 57.1% | 14 | 6.2% | | 18 | 8.0% | 194 | 85.8% |
| BMI unknown | 77 | 19.4% | 2 | 2.6% | | 5 | 6.5% | 70 | 90.9% |

| | Adults (n = 125) | | Solved (n = 5) | | Permutation Test (p-value) | Possibly solved (n = 10) | | Unsolved (n = 110) | |
|---|---|---|---|---|---|---|---|---|---|
| **All Adults (n, %)** | 125 | 100% | 5 | 4.0% | | 10 | 8.0% | 110 | 88.0% |
| **Sex (n, %)** | | | | | | | | | |
| Male | 59 | 47.2% | 2 | 3.4% | | 8 | 13.6% | 49 | 83.1% |
| Female | 66 | 52.8% | 3 | 4.5% | 0.552 | 2 | 3.0% | 61 | 92.4% |
| **Age (years)** | | | | | | | | | |
| Mean, Median (years) | 36.2, 34.1 | | 31.0, 26.1 | | | 37.5, 36.9 | | 36.3, 34.0 | |
| Range (years) | 18 – 79.8 | | 18.1 – 50.2 | | | 23.7 – 57.6 | | 18 – 79.8 | |
| **Other Symptoms (n, %)** | | | | | | | | | |
| Additional DD/ID/D | 78 | 62.4% | 3 | 3.8% | | 6 | 7.7% | 69 | 88.5% |
| No DD/ID/D | 47 | 37.6% | 2 | 4.3% | 0.623 | 4 | 8.5% | 41 | 87.2% |
| **Severity of Obesity (n, %)** | | | | | | | | | |
| Milder Obesity | 40 | 32.0% | 3 | 7.5% | 0.308 | 6 | 15.0% | 31 | 77.5% |
| Severe Obesity | 40 | 32.0% | 1 | 2.5% | | 2 | 5.0% | 37 | 92.5% |
| BMI unknown | 45 | 36.0% | 1 | 2.2% | | 2 | 4.4% | 42 | 93.3% |

Supplementary Table 4 - PanelApps

**ALL SOLVED CASES:**

| Number of Patients | Patients IDs | Gene / Locus | Full Gene name | Included in combined panel (PanelApp AUS + UK)? |
|---|---|---|---|---|
| 8 | 8 – 15 | *MC4R* | Melanocortin 4 receptor | yes |
| 3 | 16 – 18 | *PHIP* | Pleckstrin homology domain-interacting protein | yes |
| 3 | 22 – 24 | *SRRM2* | Serine/arginine repetitive matrix protein 2 | no |
| 3 | 9[a], 27, 28 | 16p11.2, proximal | | no |
| 2 | 29, 30 | 16p11.2, distal | | yes |
| 2 | 1, 2 | *ADNP* | Activity-dependent neuroprotector homeobox | no |
| 2 | 19, 20 | *RAI1* | Retinoic acid-induced gene 1 | no |
| 1 | 3 | *ALMS1* | Alstrom syndrome 1 | yes |
| 1 | 4 | *CREBBP* | CREB-binding protein | no |
| 1 | 5 | *GNAS* | eotide-binding protein,alpha stimulating activity | yes |
| 1 | 6 | *LEPR* | Leptin receptor | yes |
| 1 | 7 | *MAGEL2* | MAGE-like 2 | yes |
| 1 | 21 | *SPG11* | Spastic paraplegia 11 | no |
| 1 | 25 | *TRIP12* | Thyroid hormone receptor interactor 12 | no |
| 1 | 26 | 2p25.3 (*MYT1L*) | (Myelin transcription factor 1-like) | yes |
| | | | | **PATIENT SUM: 18/30 = 60.0%** |

*[a]: patient both with MC4R-variant and proximal 16p11.2 deletion*

**NON-SYNDROMIC:**

| Number of Patients | Patients IDs | Gene / Locus | Included in combined panel (PanelApp AUS + UK)? |
|---|---|---|---|
| 6 | 8 – 10, 12, 13, 15 | *MC4R* | yes |
| 2 | 29, 30 | 16p11.2, distal | yes |
| 1 | 9[a] | 16p11.2, proximal | no |
| 1 | 24 | *SRRM2* | no |
| | | | **PATIENT SUM: 8/9 = 88.9%** |

*[a]: patient both with MC4R-variant and proximal 16p11.2 deletion*

**SYNDROMIC:**

| Number of Patients | Patients IDs | Gene / Locus | Included in combined panel (PanelApp AUS + UK)? |
|---|---|---|---|
| 3 | 16 – 18 | *PHIP* | yes |
| 2 | 1, 2 | *ADNP* | no |
| 2 | 11, 14 | *MC4R* | yes |
| 2 | 19, 20 | *RAI1* | no |
| 2 | 22, 23 | *SRRM2* | no |
| 2 | 27, 28 | 16p11.2, proximal | no |
| 1 | 3 | *ALMS1* | yes |
| 1 | 4 | *CREBBP* | no |
| 1 | 5 | *GNAS* | yes |
| 1 | 6 | *LEPR* | yes |
| 1 | 7 | *MAGEL2* | yes |
| 1 | 21 | *SPG11* | no |
| 1 | 25 | *TRIP12* | no |
| 1 | 26 | 2p25.3 (*MYT1L*) | yes |
| | | | **PATIENT SUM: 10/21 = 47.6%** |
